# The Immunoglobulin G Glycome: A Modifiable Biomarker and Functional Effector of Aging, Disease, and Mortality

**DOI:** 10.64898/2026.04.21.26351390

**Authors:** Anika Mijakovac, Elena Butz, Frano Vučković, Azra Frkatović Hodžić, Borna Rapčan, Domagoj Kifer, Helena Deriš, Barbara Radovani Trbojević, Fran Lukšić, Ana Cindrić, Ivan Gudelj, Nina Šimunić Briški, Goran Josipović, Zehra Stara Yuksel, Jasmina Ćatić, Fran Šaler, Janko Szavits-Nossan, Charlotte R H Hedin, Jelena Šimunović, Iwona Borošak, Jasminka Krištić, Sara Monteiro-Martins, Tea Pribić, Maja Hanić, Maja Pučić-Baković, Irena Trbojević-Akmačić, Tamara Štambuk, Jerko Štambuk, Marina Martinić Kavur, Matko Fančović, Ana Cvetko, Marija Pezer, Ozren Polašek, Olga Gornik, Dobri Kiprov, Eric Verdin, Brad Younggren, Louise Newson, Cristina Menni, Claire J Steves, Tim D Spector, Jonas Halfvarson, Ewa Pocheć, Marta Szewczyk, Mirjana Turkalj, James F. Wilson, Marta E. Alarcon-Riquelme, Konrad Aden, Philip Rosenstiel, Andre Franke, Norbert Frey, Stefan Schreiber, Ellen E Blaak, Anna Köttgen, Vlatka Zoldoš, Dragan Primorac, Damir Marjanović, Ulla T Schultheiss, Wei Wang, Gordan Lauc

**Author notes:** These authors contributed equally (Anika Mijakovac, Elena Butz, Frano Vučković).

## Abstract

Glycosylation is a key structural modification of immunoglobulin G (IgG) that modulates its effector functions and has multiple roles in balancing inflammation. Altered IgG glycosylation has been reported in many diseases, often years before clinical manifestation, suggesting its causal role and biomarker potential. Here, we analyzed IgG glycome composition in 20,405 individuals from 42 different studies processed at the Genos Glycoscience Research Laboratory between 2008 and 2025. Across nearly all diseases, specific IgG glycome profiles reflected accelerated biological aging. Accelerated glycan aging was strongly associated with increased risk of all-cause mortality, independent of established clinical risk factors and potential confounders. Moreover, interventions known to reduce mortality risk, including hormone replacement therapy, therapeutic plasma exchange and caloric restriction, were associated with reversal of glycan aging. Given their role in modulating low-grade systemic inflammation, IgG glycans may represent a functional link between chronic inflammation, aging, disease susceptibility and all-cause mortality.

## Introduction

Aging represents the progressive loss of physiological integrity that ultimately leads to increased disease susceptibility and mortality. However, individuals vary widely in their ability to maintain function despite accumulating molecular and cellular damage, a property termed biological resilience, which is emerging as a key determinant of healthy aging^1^. A central process in aging is chronic inflammation, which is now recognized as one of the integrative aging hallmarks^2^. Unlike primary or antagonistic hallmarks, chronic inflammation is a systemic transducer of aging, that integrates and amplifies upstream damage and translates it into organism-wide dysfunction. It is, therefore, considered both a feature and a mechanistic driver of aging^3,4^. This dual role explains why chronic inflammation is observed not only in aging but also across a wide range of diseases, with its role in pathology cited as one of the major medical discoveries of the past two decades^5^. The ability to measure chronic inflammation may thus provide an opportunity to monitor biological aging and predict disease risk and progression. Glycans attached to the central protein of the immune system, immunoglobulin G (IgG), offer a unique possibility to quantify chronic inflammation. The importance of glycans in biology is increasingly recognized, with their structures shaped by genetics, epigenetics, and environmental factors, making them powerful integrators of complex biological systems^6,7^. These complex carbohydrates are found on most eukaryotic proteins, providing them with important structural and functional properties. Glycans, bound to the Fc region of IgG, directly affect the binding affinity of this antibody to downstream immune molecules. The composition of IgG glycans fine-tunes immune activation and resolution, thereby regulating inflammatory responses^7,8^. Over a decade ago, we reported that changes in glycans attached to IgG are a prominent feature of aging, and these changes have the potential to significantly affect inflammation and age-related diseases^9^. Based on these findings, we developed a marker based entirely on IgG glycans and demonstrated that it associates with biological age^9^. More recently, a multi-omics study of 6,304 molecular traits revealed that nine of the twenty strongest associations with chronological age were observed for glycans attached to immunoglobulins^10^. To date, numerous studies have shown that the IgG glycome changes in both aging and various diseases^8,11^. However, direct comparisons between studies and conditions remain difficult because analytical methods, data processing, and reporting standards differ between laboratories. This heterogeneity has somewhat limited the ability to draw generalizable conclusions about shared versus disease-specific patterns of IgG glycosylation. In this study, we address this gap by leveraging more than 15 years of glycomics research conducted at the Genos Glycoscience Research Laboratory. Between 2008 and 2025, we analyzed IgG glycans in almost 20,000 samples from 37 independent case-control studies, patient and population cohorts, covering 19 diseases and four aging studies. This large, systematically processed dataset allowed us to characterize IgG glycome alterations across aging and disease within a standardized framework. We further assessed the potential of IgG glycans as biomarkers of biological aging and mortality risk and explored their responsiveness to interventions.

## Results

We analyzed data from 37 different studies encompassing various diseases and aging (Supplementary Table 1). Among these, 33 studies including dedicated case control as well as observational studies that contained participants with the diseases of interest were used to investigate 19 diseases categorized into three distinct groups, described into more detail below. To improve clarity in the figures and main text, all statistical results, including p-values, are provided in the Supplementary Table 2. Also, Table 1 and Supplementary Table 1 provide an overview of all the studies and their respective categories, design, sample size and basic demographics. The first group included auto- and alloimmune diseases: systemic lupus erythematosus (SLE; four studies: SLE1, SLE2, SLE3, and SLE4), ulcerative colitis (UC; four studies: UC1, UC2, UC3, and UC4), Crohn’s disease (CD; four studies: CD1, CD2, CD3, and CD4), rheumatoid arthritis (RA; three studies: RA1, RA2 and RA3), Hashimoto’s thyroiditis (HT; one study), multiple sclerosis (MS; one study: MS1), psoriatic arthritis (PsA; one study: PsA1), Sjögren’s disease (SjD; one study: SjD1), systemic sclerosis (SSc; one study: SSc1) and graft-versus-host disease (GVHD; two studies: GVHD1 and GVHD2). The second group covered cardiometabolic diseases, with data from cardiovascular disease (CVD) and diabetes mellitus (DM). For the CVD and DM analysis, a subset of participants with prevalent CVD (N = 1460) or DM (N = 1730) as a comorbid condition from the larger German Chronic Kidney Disease cohort (GCKD) of 5217 participants diagnosed with CKD were examined. To investigate cardiometabolic diseases more closely three distinct studies representing CVD subgroups: atrial fibrillation (AF), coronary artery disease (CAD) and heart failure (HF) and one type 2 diabetes (T2D) study were also analyzed. The third group consisted of other diseases and included chronic obstructive pulmonary disease (COPD; two studies, COPD1 and COPD2), allergic sensitization to the most common inhalant and nutritional allergens (AS; two studies, AS1 and AS2) and one colorectal carcinoma (CRC) study. Additionally, four population cohorts were analyzed to quantify the effect of older age on IgG glycans: Vis (Aging1), Korčula (Aging2), ORCADES (Aging3) and TwinsUK (Aging4). Among these, the GCKD and Vis cohorts contained long-term survival data and were used for all-cause mortality analysis as a discovery and a replication cohort, respectively. To assess the effect of physiological changes on IgG glycome composition, e.g. menopause and obesity, data from Vis and Korčula cohorts was analyzed. In addition to described cross-sectional studies, three single arm longitudinal studies were conducted to assess the effect of different interventions: hormone replacement therapy (HRT), therapeutic plasma exchange (TPE) and caloric restriction.

**Table 1.**
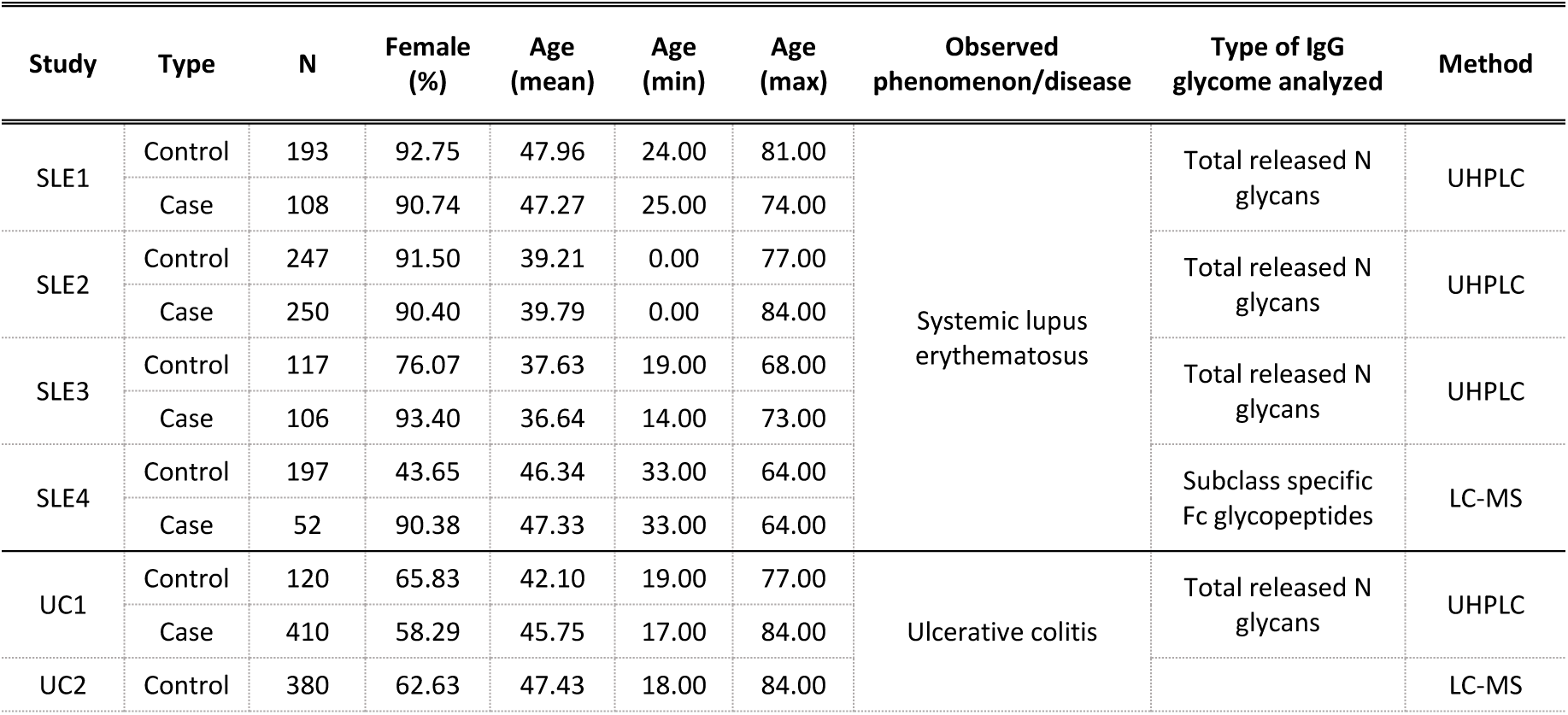
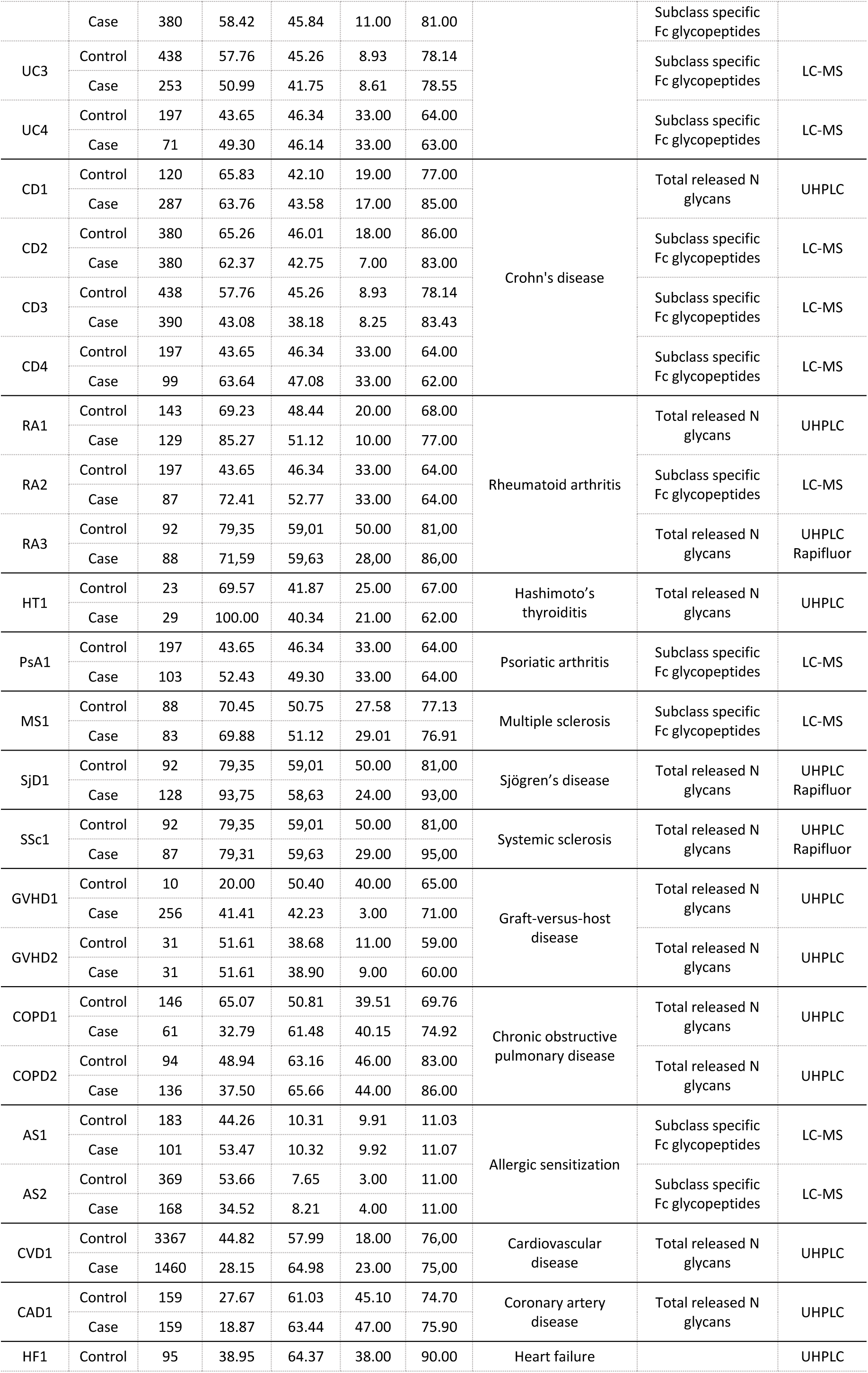
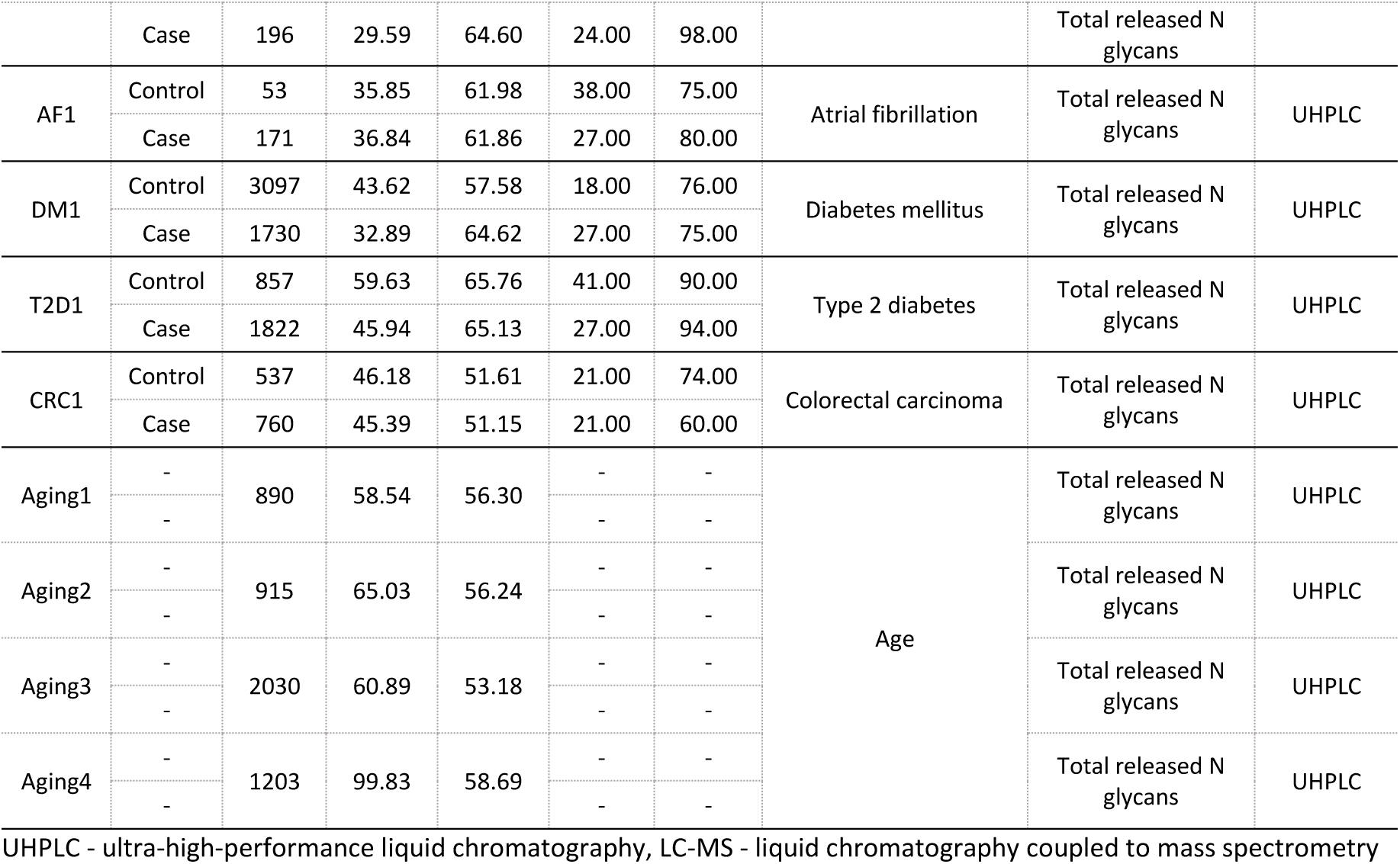
Basic characteristics of the included cross-sectional studies. This table summarizes descriptive characteristics of the included study populations including study name, number of participants, percentage of female participants, age distribution (mean, minimum and maximum), studied trait (phenomenon or disease) and the analytical methods used for IgG glycan analysis.

### IgG glycans lacking galactose are higher in disease and aging

IgG glycans can be classified based on the number of terminal galactose residues they contain: agalactosylated (G0), monogalactosylated (G1), and digalactosylated (G2). Among these, the levels of galactose-lacking G0 glycans were consistently higher amongst cases as compared to their respective controls across most of the diseases analyzed and also increased with chronological age (Figure 1A). This observation reinforces the long-standing paradigm that G0 IgG glycans increase in conditions characterized by a prominent inflammatory component. This high level of G0 glycans was particularly pronounced in autoimmune diseases, where 18 out of 20 studies exhibited a statistically significant difference compared to the respective controls (Figure 1A; Supplementary Table 2). The most comprehensive datasets originate from SLE, UC, and CD, where analyses of four independent studies per condition consistently demonstrated elevated IgG G0 glycan levels in affected individuals relative to unaffected controls. Similar patterns were observed in patients with RA, the most extensively studied autoimmune disease with regards to IgG glycosylation. Also, elevated levels of G0 IgG were found in PsA, SjD and SSc. Non-significant upward trends were observed in both HT and MS compared to their respective controls. Of note, HT and MS were the smallest autoimmune studies analyzed (29 cases and 23 controls for HT; 83 cases and 88 controls for MS), which likely limited statistical power (Table 1). The observed patterns of G0 IgG glycans in autoimmune diseases are in line with findings from independent studies reported from different research groups^11^. In contrast, data on IgG glycosylation in alloimmune disorders remains limited with only a few studies available on fetal neonatal alloimmune thrombocytopenia (FNAIT)^12,13^. Here, we analyzed two studies of GVHD and observed a trend toward higher levels of IgG G0 glycans in patients compared to their respective controls, though the difference was not statistically significant. A similar trend was observed in COPD, another disease with a significant inflammatory component. Unexpectedly, no differences in G0 IgG glycans were observed between AS participants and their respective controls. However, both AS studies were performed in children aged 3 to 11 years, a critical distinction, as pediatric populations may exhibit different IgG glycome dynamics compared to adults, potentially explaining the apparent stability of IgG glycome in this group^11^. A considerably higher level of G0 IgG glycans when compared to the respective controls was observed in DM but also specifically in T2D, an age-related diabetes type with a strong inflammatory component. The same pattern was observed in all four aging cohorts, corroborating previous findings of a strong association of G0 IgG glycans with increasing age^9^. To further investigate the association of IgG glycans with age-related disease we analyzed the changes of IgG glycans in CVD and observed higher levels of G0 glycans in CVD when compared to the respective controls. Considering the heterogeneity of CVD, we also analyzed IgG glycans in three distinct CVD subgroups: CAD, HF and AF. Interestingly, HF was the only CVD subgroup where we observed a strong positive association of disease status with G0 glycans. The association was much weaker for CAD and a converse trend was observed for AF, but none were statistically significant. Strong positive association with G0 IgG glycans was observed in CRC, the only cancer study included. Comparable patterns have been reported by other groups in both solid and liquid tumors, suggesting that this glycosylation pattern may be common in different malignancies^11^.

**Figure 1.**
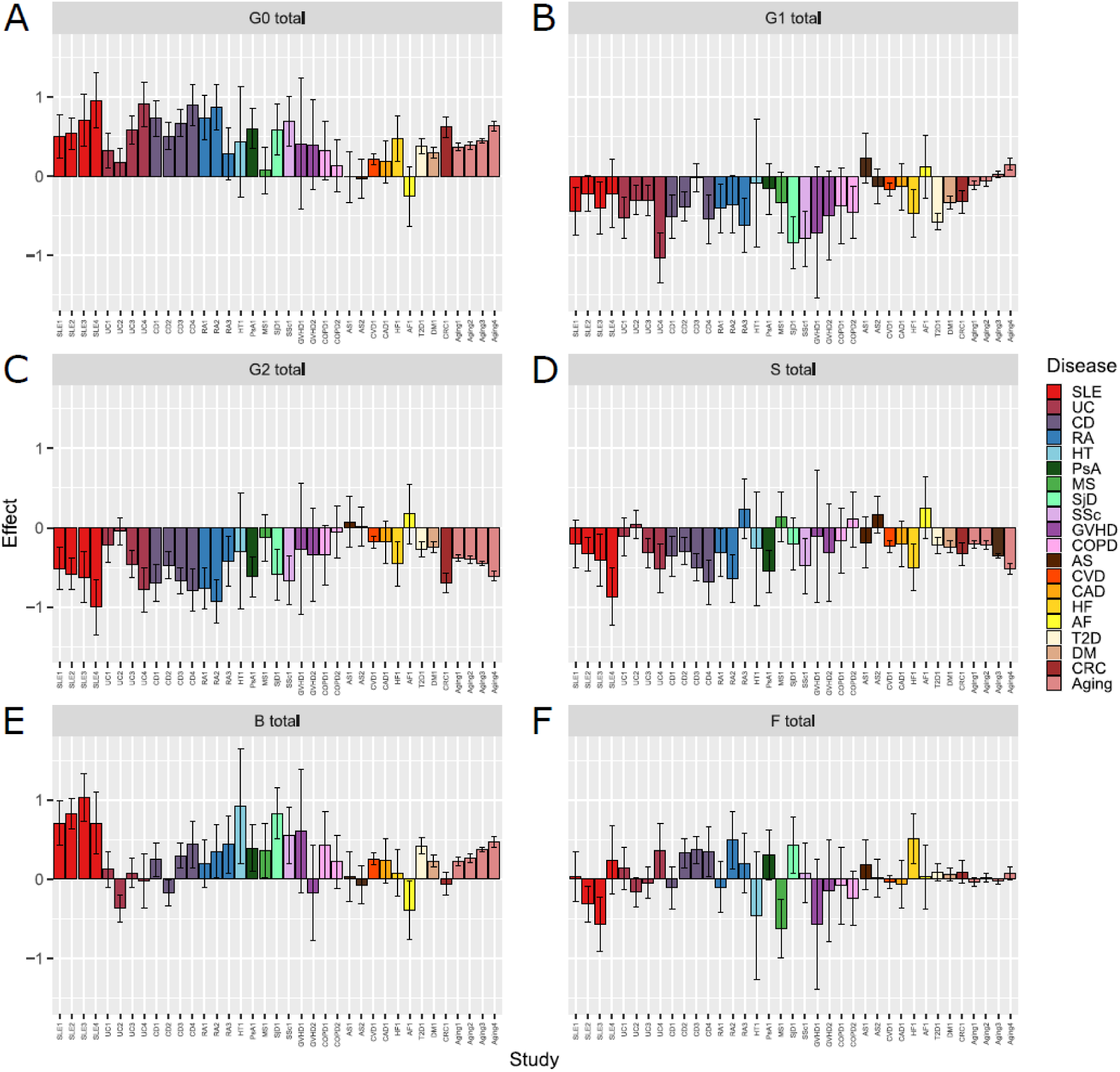
IgG glycome differs by disease status and age. Derived IgG glycosylation traits were calculated from glycan/glycopeptide measurements obtained using UHPLC or LC-MS analyses. The following traits are shown: **A** - total agalactosylated glycans (G0); **B** - total monogalactosylated glycans (G1); **C** - total digalactosylated glycans (G2); **D** - total sialylated glycans (S); **E** - total glycans with bisecting GlcNAc (B); and **F** - total fucosylated glycans (F), as previously described ^59^. The X axis shows the individual studies representing the status of different diseases. The Y axis shows regression coefficients (β) obtained from linear models, representing the effect of disease status or chronological age on the abundance of the respective glycan trait. For disease models, β represents the mean difference between cases and controls (in standard deviation units). For aging cohorts, β represents the change in glycan trait per 10-year increase in chronological age (SD units per decade). Models assessing disease status were adjusted for age and sex, whereas models assessing chronological age were adjusted for sex only. All glycan traits were transformed to a standard normal distribution (mean = 0, SD = 1) using inverse rank-based normal transformation, enabling direct comparison of effect sizes across traits and studies.

### IgG galactosylation is lower in disease and aging with notable differences between monogalactosylation (G1) and digalactosylation (G2)

The levels of IgG glycoforms containing terminal galactose residues were lower in older individuals and most of the analyzed diseases as compared to respective controls (Supplementary Figure 1). However, a more detailed examination revealed notable differences between monogalactosylated (G1) and digalactosylated (G2) IgG glycans, with the overall larger effect consistently observed for G2 IgG glycans (Figure 1B and Figure 1C). Among autoimmune diseases, G2 glycans were substantially lower than G1 in several studies, including SLE, CD, RA, and PsA. Furthermore, in PsA, CD3 and SLE3 no associations were detected between disease and G1 glycan levels. Conversely, in patients with UC, MS, SjD and SSc a much larger effect was observed for G1 IgG glycans, except in the UC3 study. Neither G1 nor G2 IgG glycan levels differ in HT and AS compared to their respective controls. A trend towards lower overall IgG galactosylation was observed in both GVHD and COPD cases relative to their respective control groups, although these changes did not always reach statistical significance. Larger effect was also observed for G1 than G2 IgG glycans in DM and specifically in T2D. In CRC patients relative to their respective controls, however, the lower levels of G2 IgG glycans were considerably more pronounced compared to G1. Of note, the most prominent difference in the effect size for G1 and G2 IgG glycans was detected in the aging cohorts. G1 glycans were relatively stable, showing no change in the Aging3 cohort, a slight age-related decrease in both the Aging1 and Aging2 cohorts, and a modest age-related increase in the Aging4 cohort. In contrast, G2 glycans demonstrated a strong and consistent decrease across all four aging cohorts. On the other hand, in CVD overall and specifically in patients with HF compared to controls, significantly lower level of both G1 and G2 glycans were observed. In patients with AF and CAD both G1 and G2 IgG glycans remained stable. Even though global IgG galactosylation follows a similar pattern of change in both diseases and aging, distinguishing between IgG glycans with one or two galactose residues revealed markedly different effects. These findings suggest that G1 and G2 glycans may serve as distinct biomarkers, with aging being uniquely characterized by G1 stability and G2 decline, while many disease states exhibit concurrent reduction in both when compared with controls.

### Sialylated IgG glycans are lower in disease and aging

In most of the analyzed studies, sialylated IgG glycans were significantly lower in patients than in the respective controls and were lower in older individuals. However, it must be emphasized that galactosylated IgG acts as a substrate in the process of sialylation, so the dynamics of IgG sialylation are inherently tied to IgG galactosylation. This strong positive correlation between galactosylated and sialylated IgG glycans is reflected in our dataset, where sialylation patterns largely mirror those of digalactosylated (G2) IgG structures across most studies (Figure 1C and Figure 1D). Sialylated IgG glycans were significantly lower in SLE, CD, UC, RA, PsA and SSc than in the corresponding control populations, with no change reported in SLE1, UC1, UC2 and RA3 studies. IgG sialylation also remained unchanged compared to the respective controls in HT, MS, SjD, GVHD, COPD, AS, CAD and AF. On the other hand, a prominently lower level of sialylated IgG was observed in the HF study as well as in CVD overall compared to the respective controls. The same prominent trend was observed in T2D, CRC and all four aging cohorts, demonstrating that both in age and age-related diseases the levels of IgG sialylation are lower than among the respective controls.

### Bisected IgG glycans are higher in some diseases and aging

In contrast to the extensively documented changes in IgG galactosylation over the past three decades, alterations in the levels of bisecting *N*-acetylglucosamine (GlcNAc; B) have been less frequently reported. This is partly due to technical limitations in accurately detecting bisected structures using older analytical methods. In this study, we observed an overall higher level of bisected IgG glycans in aging and diseases compared to the respective controls, with a few exceptions (Figure 1E). The most prominently higher levels of bisected IgG glycans were observed in SLE, HT, SjD, SSc, CVD, DM and T2D cases in relation to their respective controls. Comparable effects were also found in aging, with older people having higher levels of bisecting GlcNAc-containing IgG glycans. Interestingly, in three (UC2, CD2 and AF) out of 37 analyzed studies, a significantly lower level of bisected IgG glycans was detected in cases compared to controls. These outlier findings stand in contrast to the broader pattern and may warrant further investigation to determine whether they reflect true biological variability or study-specific effects. Across all the other analyzed diseases, bisected IgG glycans were higher or remained stable compared to controls.

### IgG glycans with core fucose remain stable in most diseases and aging

Core fucosylation is a defining feature of the IgG glycome, with approximately 95% of IgG glycans containing a core fucose residue. This modification plays a key role in modulating IgG effector function, notably by impairing antibody-dependent cellular cytotoxicity (ADCC) via reduced affinity for FcγRIIIA^14,15^. Despite its immunological relevance, changes in IgG fucosylation have rarely been reported in glycomic studies to date^16,17^. This may, in part, reflect the technical challenges associated with accurately quantifying the low-abundance afucosylated species using earlier analytical platforms. Our data demonstrates that core fucosylation levels did not show pronounced differences in most studies (Figure 1F). The most notable exception was observed in MS patients, where a significantly lower level of IgG core fucosylation was detected compared to the respective controls. The same was observed in SLE2 and SLE3 and was not replicated in the other two analyzed SLE studies. Higher levels of core fucosylated IgG glycans were observed in some CD, UC, RA and SjD cases compared to their respective controls but this was not replicated in all the analyzed studies. From all the analyzed cardiometabolic diseases, a significantly higher level of core fucosylation was observed only in HF compared to the respective controls. IgG core fucosylation remained stable with aging apart from Aging4 study in which slightly higher levels were observed. These intra- and inter-disease discrepancies may stem from differences in cohort size, disease severity, or study design, as well as potential variability in analytical sensitivity. Given the relatively small effect sizes observed, detecting significant shifts in IgG fucosylation requires large, well-powered cohorts. Future studies, including those employing high-sensitivity glycoprofiling techniques, will be needed to resolve these inconsistencies and further evaluate the biological significance of fucosylation in disease.

### Glycan aging is accelerated in disease

IgG glycosylation strongly associates with age in all four analyzed population cohorts as shown in Figure 1, corroborating previous findings^18^. The most pronounced and consistent age-related changes were observed in the agalactosylated (G0), digalactosylated (G2) and sialylated (S) IgG glycan structures (Figure 1A, Figure 1C and Figure 1D), which formed the basis for the development of the glycan age^9^. While this biomarker can predict chronological age with reasonable accuracy, its greater significance lies in its association with a broad range of biochemical and physiological factors indicative of biological aging. To investigate the rate of biological aging across various pathological conditions, we calculated glycan age in 33 studies encompassing 19 distinct diseases. We did the same in the four population cohorts to confirm the association of glycan age and chronological age. Glycan age was calculated for each dataset separately by implementing a general regression model where G0, G2 and S were used as predictors, chronological age was used as the response variable, and model prediction was defined as glycan age. In the four population cohorts (Aging1-4), instead of case/control status, chronological age (expressed in decades) was included in regression model as a predictor. Strikingly, glycan age was accelerated in nearly all examined disease studies relative to the respective controls (Figure 2A). In patients with autoimmune diseases, glycan age was, on average, 2.1 years higher than in matched healthy controls. This increase did not reach statistical significance in one SLE, one UC study and in SjD. The same is true for MS and HT, but it must be emphasized that these studies were of the smallest scale limiting the interpretability of the results. A trend toward glycan age acceleration was observed in both GVHD studies - the only alloimmune disease included in the analysis. Allergic sensitization, another immune-mediated condition showed no association with glycan age. Notably, both allergic sensitization studies consisted exclusively of young children, and no adult participants were included. This may explain the lack of association, especially given previous reports that describe inconsistent changes in the IgG glycome in pediatric populations, with no clear age-dependent trends^11^. Considering the established clinical relevance of IgG in allergic responses, further investigation of allergic sensitization in adult populations would be of high value, although this lies beyond the scope of the present study. Analysis of age-related diseases revealed consistent evidence of higher glycan age across most studies. In both COPD studies, glycan age was higher than in the respective controls, but the results were not statistically significant. When examining CVD as one condition, glycan age was higher compared to non-CVD individuals. A more detailed analysis of CVD subtypes revealed significant glycan age acceleration specifically in HF, whereas no significant changes were observed in CAD or AF, where glycan age remained unchanged relative to their respective controls. Notably, pronounced glycan age acceleration was observed in CRC as well as in both DM and T2D studies in reference to their respective controls, highlighting the sensitivity of this biomarker in capturing biological aging across a range of age-associated and inflammation-driven conditions.

**Figure 2.**
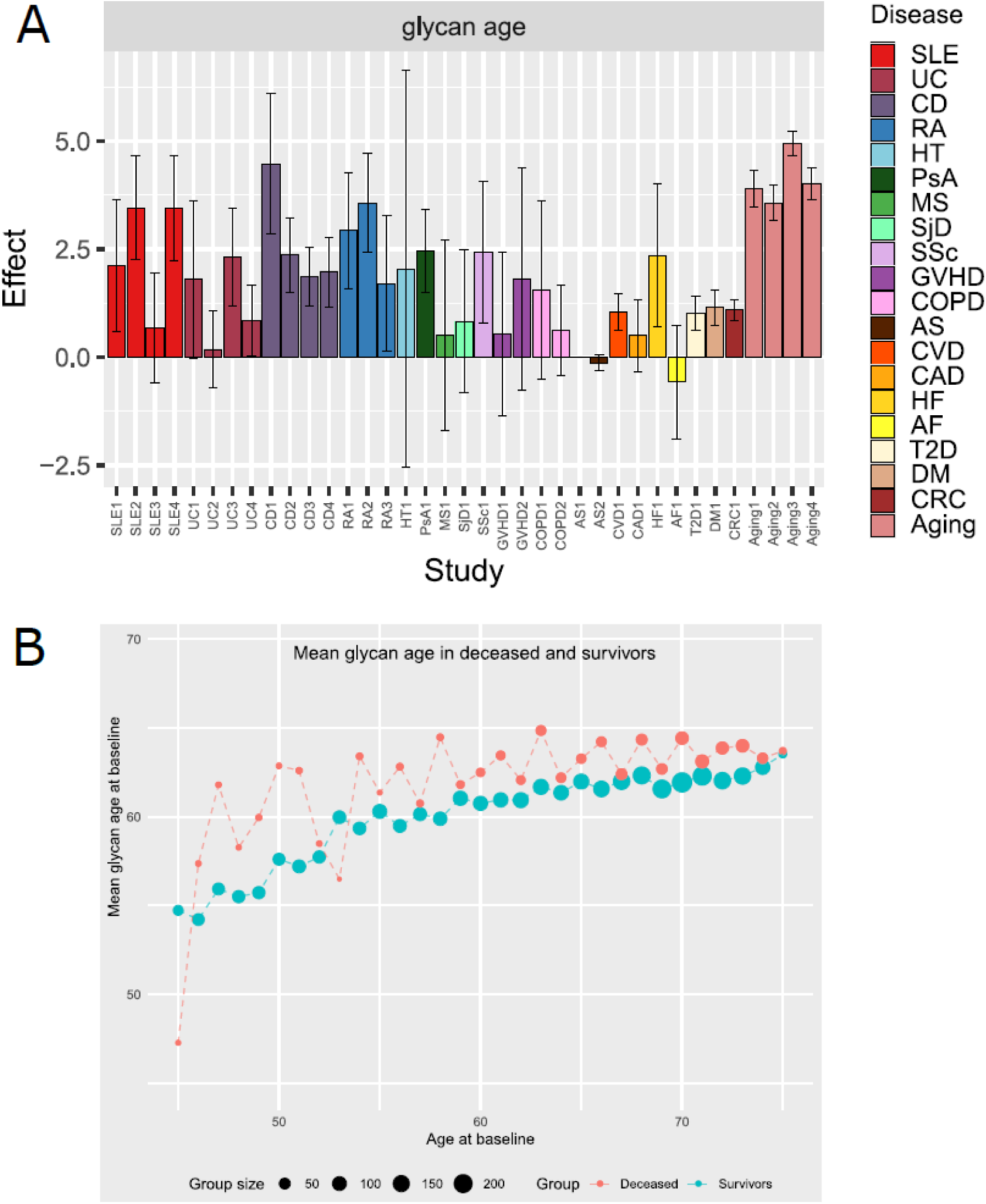
Glycan aging is higher in disease and associated with all-cause mortality. **A** –Glycan age was calculated using general regression model where G0, G2 and S were used as predictors, chronological age as response variable, and the model prediction was defined as glycan age. The X axis shows the individual studies representing the status of different diseases. The Y axis shows regression coefficients (β) obtained from linear models, representing the effect of disease status or chronological age on the glycan age. For disease models, β represents the mean difference in glycan age between cases and controls. For aging cohorts, β represents the change in glycan age per 10-year increase in chronological age. Models assessing disease status were adjusted for age and sex, whereas models assessing chronological age were adjusted for sex only. **B** – Y axis shows mean glycan age at the baseline visit for participants of the GCKD study. X axis shows chronological age (in years) at the baseline visit of the GCKD study (ages ≥ 45 shown due to sparse data at younger ages). Blue dots represent participants of the GCKD study that did not die during the follow-up period of 8.5 years. Red dots represent the participants of the GCKD study that died during the follow-up period due to any cause. All analyses were carried out in the GCKD study, used as a discovery cohort.

### Accelerated glycan aging is associated with higher all-cause mortality risk, independent of traditional clinical risk factors

To examine the association between accelerated glycan aging and all-cause mortality, we applied Cox proportional hazards regression in the GCKD cohort. In the 4,827 participants (60% male), 840 deaths occurred over a median follow-up of 8.5 years (data freeze: 2024/11/21), with men accounting for a disproportionate share (639 of 840 deaths). In a model adjusted for chronological age and sex (M1), each additional year of glycan age was associated with a 10% increase in the hazard of all-cause mortality (HR 1.10, 95% CI 1.08–1.12), as shown in Supplementary Tables 3 and 4. This association remained significant after further adjustment for established mortality risk factors (body mass index (BMI), smoking status, systolic blood pressure, low-density lipoprotein (LDL) cholesterol, C reactive protein (CRP), and diabetes) and kidney function markers (eGFR and uACR). In this fully adjusted model (M4), each year of additional glycan age was associated with a 5% increase in the mortality hazard (HR 1.05, 95% CI 1.03-1.06). To replicate these findings, we repeated the analysis in the population-based Vis cohort, which included 796 individuals and 94 recorded deaths over a 10-year follow-up period. After adjusting for chronological age, sex, and the same set of known mortality risk factors (except kidney function markers and CRP; M2), each additional year of glycan age was associated with a 3% increase in the hazard of all-cause mortality (HR 1.03, 95% CI 1.00–1.06) as shown in Supplementary Table 5. We were not able to replicate the results from the discovery cohort in the Vis cohort using the fully adjusted model (M3) that besides all the established mortality risk factors also contained CRP. Finally, we compared glycan ages between participants of the same chronological age whose death had been observed (cases) or not observed (controls) during follow- up. In the GCKD study, a subgroup of participants aged 45–64 years at baseline who died during follow-up (N = 255) had higher mean glycan ages compared to surviving participants (N = 1743) within the same age group (62 ± 5 vs. 59 ± 6; *P* = 4.7 × 10⁻^17^, two-sided t-test) as shown in Figure 2B and Supplementary Table 6. This was also replicated in the Vis cohort as shown in Supplementary Figure 2 and Supplementary Table 6. A similar difference was observed in the 65–75 age group, with higher glycan ages among deceased participants (N = 577) compared to survivors (N = 1,674; mean glycan age 64 ± 4 vs. 62 ± 5; *P* = 1.9 × 10⁻¹^3^), but we weren’t able to replicate this finding in the Vis cohort (Supplementary Table 6).

### Glycan aging is accelerated in menopause and rejuvenated by hormone replacement therapy

We demonstrated that glycan age, a biomarker derived from IgG glycans, is accelerated in various diseases relative to their respective controls and this acceleration in glycan age is significantly associated with mortality risk in these populations. Other studies have also shown that IgG glycans dynamically change in response to physiological alterations. One such major physiological shift is menopause, during which the IgG glycome undergoes marked and consistent alterations^19^. To examine the impact of menopause on glycan aging, we compared glycan age of menopausal and premenopausal women matched for chronological age, using data from two Croatian population cohorts, Vis (N, matched = 84) and Korčula (N, matched = 106) described in Supplementary Table 7. We found that premenopausal women exhibited, on average, a glycan age that was 8 years younger than their menopausal counterparts (95% CI 5.7 – 10, *P* < 0.001) as seen in Supplementary Figure 3. These findings support the hypothesis that menopause triggers a sharp acceleration in biological aging likely mediated by hormonal imbalances, specifically estradiol, progesterone and testosterone depletion^20,21^. To explore whether treatment with body identical hormone replacement therapy (HRT) might mitigate this acceleration, we monitored 19 healthy perimenopausal/menopausal women who underwent HRT at the Newson Clinic (Supplementary Tables 8 and 9). Glycan age was assessed multiple times over the course of 15 months with variable sampling frequencies across participants. We observed a highly significant and consistent reduction in glycan age over time, with an average decrease of 0.12 glycan years per month of HRT (*P* = 5.76 × 10⁻⁸; Figure 3A, Supplementary Table 10). This rejuvenating effect emphasizes the potential of body identical HRT to reverse menopause-associated biological aging at the glycomic level.

**Figure 3.**
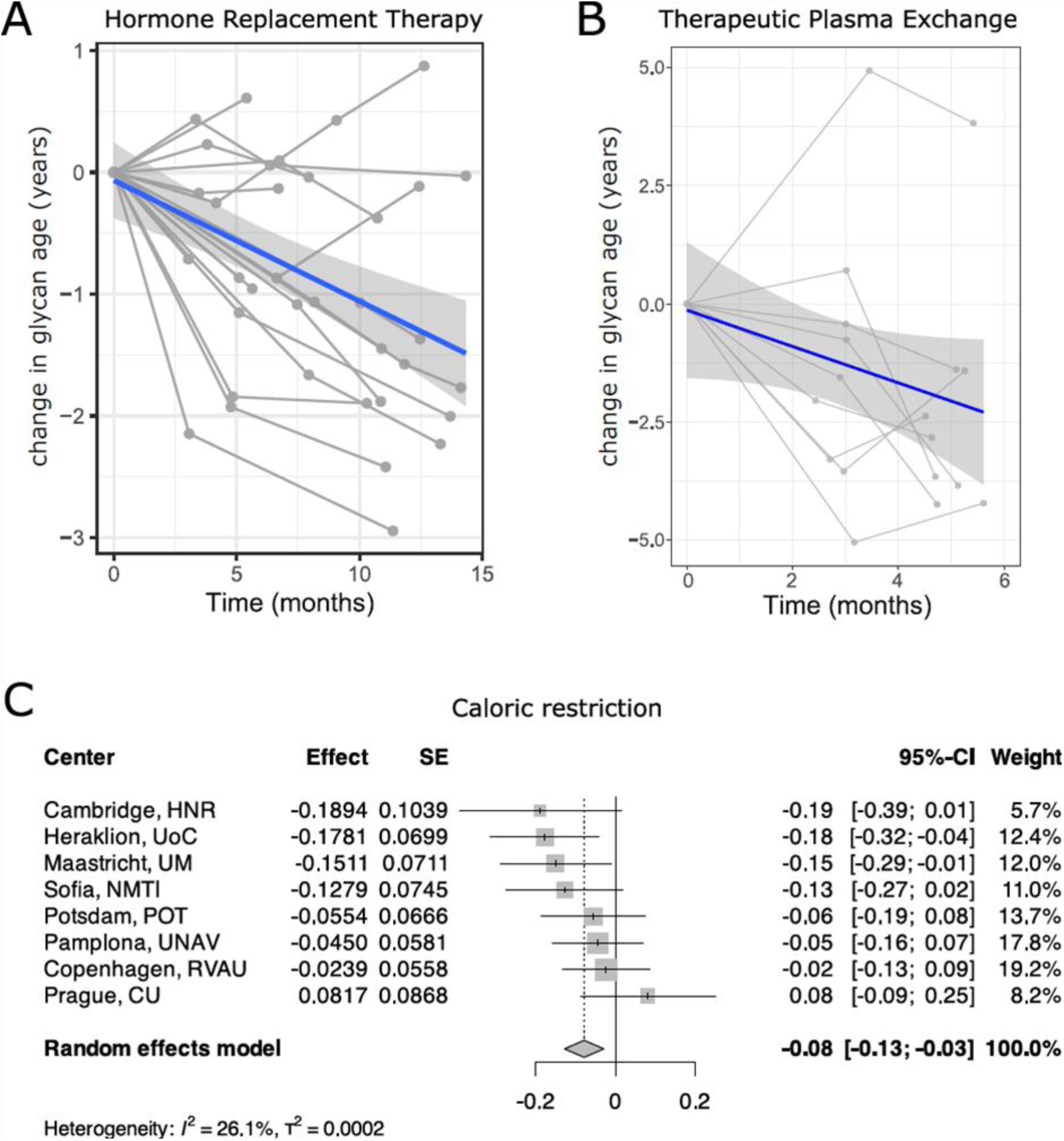
Geroprotective interventions deaccelerate biological age as measured by IgG glycans. **A** - Changes in glycan age in women receiving HRT over the course of 15 months and **B -** Changes in glycan age from baseline after TPE therapy over 6 months. Change in glycan age was calculated as the difference between glycan age at each measurement and baseline. Blue line represents a linear regression fit modelling the relationship between time (in months) and delta glycan age. The shaded area indicates the 95% CI. **C** - Effect of caloric restriction and consequent weight loss on glycan age after 8 weeks. Changes in glycan age obtained through a meta-analysis across all centers are shown. SE, standard error; 95% CI, 95% confidence interval.

### Therapeutic plasma exchange deaccelerates glycan aging

Therapeutic plasma exchange (TPE), long established as a treatment for various diseases, even as a first line option for conditions such as Myasthenia gravis and Guillain-Barre syndrome is now gaining renewed attention in the context of anti-aging and longevity^22–24^. To investigate the effect of TPE on biological aging as captured by the IgG glycome, we longitudinally profiled IgG glycans in 9 individuals who underwent monthly TPE sessions over a 6-month period (Supplementary Tables 9 and 11). We observed a decline in glycan age, with an average reduction of 0.4 glycan years per month of TPE treatment. (*P* = 0.0245); Figure 3B, Supplementary Table 10). This finding suggests that repeated plasma exchange may exert a rejuvenating effect on the IgG glycome, highlighting its potential as an anti-aging intervention.

### Glycan aging accelerates in obesity but is restored with weight loss

Obesity, now increasingly recognized as a chronic systemic disease and loosely defined as a body-mass index above 30 kg/m^2^, is associated with complex hormonal and metabolic disturbances. Prior research has linked obesity to alterations in the IgG glycome, particularly a reduction in sialylation, which has been implicated in the pathophysiology of obesity-related hypertension and insulin resistance^25,26^. To investigate whether higher BMI influences glycan aging, we analyzed data from two Croatian population cohorts: Korčula (N = 915) and Vis (N = 886) described in Supplementary Table 12. Our analysis revealed a significant positive association between BMI and glycan age: for each unit increase in BMI, glycan age increased by an average of 0.35 years (95% CI 0.24 – 0.46 years; *P* < 0.001; Supplementary Figure 4, Supplementary Table 13) keeping the age constant at 56 years (dataset mean). Interestingly, the association between BMI and glycans age was weaker in older individuals (Supplementary Table 14) suggesting that the influence of adiposity on biological aging may attenuate with age. Next, we sought to determine whether glycan aging is reversible through caloric restriction and weight loss. We calculated glycan age from samples collected at eight European centers that took part in the DIOGENES intervention study (N = 680), in which participants underwent an 800 kcal/day low-calorie diet for eight weeks^27,28^ (Supplementary Table 9). All centers reported substantial weight loss accompanied by a decreasing trend in glycan age in seven of the eight centers (Figure 3C, Supplementary Table 10, Supplementary Figure 5). Meta-analysis across sites demonstrated that an average weight loss of 11 kg resulted in a significant deacceleration of glycan age (*P* = 0.0017, Figure 3C), indicating that obesity-driven biological aging, as captured by glycan age, is reversible through caloric restriction.

## Discussion

The geroscience hypothesis proposes that a common set of interconnected biological pathways underlies the increased vulnerability to multiple diseases observed with aging, thereby driving multimorbidity^29^. Biomarkers that capture these fundamental processes can provide integrated molecular measures of biological aging and resilience even serving as surrogate end points for age-associated outcomes in clinical studies. Recently, three landmark papers from the Biomarkers of Aging Consortium outlined the conceptual and methodological criteria that define a valid biomarker of aging^30–32^. First and foremost, an aging biomarker must reflect underlying biology and be anchored in the established hallmarks of aging. Extensive research over the past decade has demonstrated that IgG glycans regulate inflammation, the most integrative hallmark of aging that underlies different pathologies^8,33^. However, to serve as valid biomarkers they must also predict health outcomes associated with aging such as morbidity, mortality and functional decline. In this study, we demonstrated that IgG glycome composition changes both with higher age and across different diseases, and that these alterations share a common pattern characterized by lower galactosylation and sialylation. By harmonizing measurements across 37 independent studies encompassing almost 20,000 individuals from diverse populations, we were able to compare, for the first time, disease-and age-associated glycomic profiles within a unified analytical framework. The scope of this unified analysis was further strengthened by the addition of 11 newly generated, unpublished datasets with more than 6000 samples extending the coverage of autoimmune and cardiometabolic studies. This unique large-scale integration revealed that most diseases recapitulate glycan signatures of aging, consistent with a shared pro-inflammatory shift. This limits the biomarker potential of IgG glycans in diagnosing individual diseases. However, traditional disease classification often fails to capture the underlying molecular pathology. Considering that IgG glycans are established functional effectors of the immune system, the changes we observed may indeed reflect shared molecular pathways across diseases. Upon closer inspection of the derived IgG glycan traits across different diseases, we also found some disease-specific changes such as the consistently higher proportion of bisection across all four SLE studies compared to the respective controls and the lack of association between IgG monogalactosylation and higher age but significant reduction of monogalactosylated IgG glycoforms in most disease subgroups. Such direct comparison of disease- and age-related glycomic profiles would not have been possible without the standardized analytical framework applied here. Further efforts to expand this framework to a larger number of diseases and cohorts will be essential to determine whether specific IgG glycans could serve as disease-specific biomarkers. To extend beyond cross-sectional associations and examine the association of IgG glycans with future events, we performed survival analysis, a gold standard to evaluate aging biomarkers as outlined by the Biomarkers of Aging Consortium^30–32^. Unlike classical mortality analysis, this approach enabled us to capture dynamic, time-to-event information, reflecting the continuous nature of biological aging. In a large, prospective CKD cohort (GCKD; n = 4,827; median follow-up 8.5 years), higher glycan age was associated with a markedly increased risk of all-cause mortality even after controlling for established risk factors including BMI, smoking, blood pressure, LDL cholesterol, CRP, prevalent diabetes mellitus, and kidney function. This was successfully replicated in the Vis cohort (n = 796; 10-year follow-up) after adjusting for all established risk factors except the inflammatory biomarker CRP further demonstrating the role of IgG glycans in inflammation. However, in the larger discovery cohort the association of glycan age with increased risk of all-cause mortality remained strong even after the inclusion of CRP. Although CRP is an established marker of systemic inflammation it predominantly reflects the acute-phase response limiting its ability to capture long-term inflammatory burden which may be more accurately reflected by IgG glycans that are less sensitive to transient inflammatory exposures. To our knowledge this is the first study demonstrating that IgG glycans not only associate with aging but are associated with incident death over time, independent of chronological age and conventional clinical risk factors. Moreover, in both the discovery and replication cohort we also demonstrated that participants who died during the follow-up period in both cohorts consistently exhibited higher baseline glycan age than their age-matched survivors. Other prospective studies also demonstrated the predictive validity of IgG glycans. For example, IgG glycome was shown to be predictive of future development of rheumatoid arthritis^34^, relapse of vasculitis^35^, and progression of ulcerative colitis and Crohn’s disease^36,37^. Large studies also revealed that it associates with cardiovascular disease risk score - American Heart Association risk score (AHA)^38^ and even predicts future cardiovascular events^39,40^. The fact that IgG glycans change years before the onset of clinical symptoms^34,38^ suggests that they may serve as an early warning signal for personalized predictive medicine. Beyond predictive capacity, an informative biomarker of aging must also be dynamic - able to capture both the acceleration of aging in adverse physiological states and the restoration of resilience following interventions known to promote longer health span. We therefore investigated whether IgG glycans could capture this bidirectional plasticity of biological aging across three mechanistically distinct yet widely recognized geroprotective interventions: hormone replacement therapy (HRT), therapeutic plasma exchange (TPE) and caloric restriction. Menopause, characterized by hormonal decline and heightened inflammatory status, was accompanied by a sharp increase in glycan age - premenopausal women were approximately eight biological years younger than their age-matched postmenopausal peers. HRT reversed this effect with an average decline of 0.12 glycan years per month of treatment. TPE, an emerging gerotherapeutic modality, reduced glycan age by an average of 0.4 years per month of treatment demonstrating, for the first time, the beneficial effect of this procedure on biological age measured using IgG glycans. Obesity, another state of chronic low-grade inflammation, was associated with accelerated glycan aging (0.35 years per BMI unit), while caloric restriction produced the opposite effect, significantly decelerating glycan aging following weight loss. These findings demonstrate that IgG glycans can also serve as personalized response biomarkers that capture the reversible component of aging biology reflecting both the loss and restoration of physiological resilience. In addition to the core criteria, biological relevance, predictive validity, and responsiveness to interventions, a robust biomarker must also be analytically robust and minimally invasive. IgG glycans have clear advantages in this regard: they can be quantified with high precision, measured from plasma or dried blood spots, and remain stable under routine storage conditions^41,42^. Another key requirement is cross-species validation which enables better understanding of the biological relevance of the biomarker and translational applicability in preclinical and clinical studies. Unfortunately, analysis of IgG glycans in various murine models yielded conflicting results potentially due to attenuated immune activation in the pathogen-free environment^43^. Conversely, a recent study found that in rat models young porcine plasma reversed biological age measured both by epigenetic clocks and IgG glycans^44^. Finding suitable models is particularly important given the fact that IgG glycans function as effectors of immune regulation. Although this study doesn’t provide causal proof that IgG glycans contribute to the risk of disease progression or death, considering their established role in the immune system this is not an unlikely hypothesis. It is conceivable that beyond being biomarkers they may emerge as therapeutic targets which will warrant rigorous experiments both *in vitro* and *in vivo.* While this study provides a comprehensive and integrative analysis of IgG glycosylation in aging and disease, several limitations must be considered. Although the diversity of the study populations represents one of the main strengths of this study, it also introduces residual heterogeneity, and some associations could partly reflect study composition rather than disease biology. Nevertheless, the consistent direction of the effects across independent datasets supports the robustness of the underlying signal. Another limitation is the size of some disease studies, such as GVHD2, HT, and MS, that were relatively small (<200 participants), and replication in larger, independent populations is warranted. In addition, autoimmune diseases are overrepresented in our dataset, reflecting their early prioritization in IgG glycosylation research due to the clear immune component of their pathology. However, chronic inflammation is now recognized as a key driver of not only autoimmune but also cardiovascular and metabolic diseases^45–47^, in which we indeed observed significant alterations of IgG glycome composition. Cancer and neurodegenerative diseases remain notably underrepresented, limiting our ability to explore their effect on IgG glycome composition. Future large-scale efforts integrating more disease groups – particularly cardiometabolic, oncological and neurodegenerative - within a harmonized analytical framework will be crucial to establish the generalizability of our findings and define disease-specific versus shared IgG glycome alterations. Our intervention studies, particularly those examining HRT and TPE, were also limited by relatively small sample sizes, a common challenge in biomarker studies involving intensive longitudinal sampling. Future work should aim to recruit larger cohorts to quantify the more nuanced effects of different interventions on IgG glycome composition. In conclusion, this study provides convincing evidence that IgG glycans can serve as promising candidates for clinically actionable biomarkers of biological aging. Their mechanistic grounding and capacity to capture both decline and recovery underscore their potential as tools for measuring resilience, and, perhaps one day, as levers to restore it.

## Materials and methods

### Study descriptions

This research integrates data from a total of 20,045 participants originating from multiple independent studies, both cross-sectional and prospective, encompassing healthy individuals and patients with a range of diseases. Participants were recruited across diverse international sites, ensuring broad demographic and clinical representation. In all studies, IgG *N*-glycome profiling was performed using standardized UHPLC, LC–MS or CGE methodologies, analyzing either total IgG or subclass-specific Fc glycopeptides according to the respective study protocols. Each study was conducted in accordance with the principles of the Declaration of Helsinki and approved by the relevant institutional or national ethics committees, with all participants providing written informed consent prior to enrolment. For glycomic datasets that have been previously published, the corresponding PMID references are provided in the Supplementary Table 1, and detailed descriptions of the study design, recruitment, and analytical procedures can be found in their original publications. In addition to these published datasets, several unpublished glycomic datasets were included in the present study to expand coverage and improve population diversity. The descriptions of these studies are provided below.

#### Heart Failure

The heart failure (HF) study consisted of 191 patients with heart failure and 95 control subjects, all recruited at University Hospital Dubrava, Zagreb, Croatia. The heart failure group had a mean age of 65 years, including 135 men (71%) and 55 women (29%). Clinical severity was distributed across New York Heart Association (NYHA) functional classes I–IV, with most patients in class II (58%) and class III (23%). Based on left ventricular ejection fraction (LVEF) (mean 39%), patients were classified as Heart Failure with Reduced Ejection Fraction (HFrEF) (n=103, 54%), (Heart Failure with Mildly Reduced Ejection Fraction) HFmrEF (n=33, 17%), and Heart Failure with Preserved Ejection Fraction (HFpEF) (n=51, 27%). Circulating N-terminal pro–B-type Natriuretic Peptide (NT-proBNP) levels were markedly elevated (mean ∼4047 pg/mL). The control group included 95 individuals (mean age 64 years), comprising 58 men (61%) and 37 women (39%), without clinical heart failure. Plasma IgG was isolated and analyzed for *N*-glycan composition using UHPLC under the same experimental conditions as for the other studies. The study was conducted in accordance with the Declaration of Helsinki and approved by the Ethics Committee of University Hospital Dubrava.

#### German Chronic Kidney Disease

The German Chronic Kidney Disease (GCKD) study is a well described^48,49^ and ongoing multi-center prospective cohort study that recruited 5,217 participants with chronic kidney disease from 169 nephrology practices as well as nine university clinics across Germany. To be eligible, patients had to have an eGFR of 30–60 mL/min/1.73m² or >60 mL/min/1.73m² with excess albuminuria. Clinical endpoints are being prospectively collected in a standardized manner, with ongoing follow-up based on hospital discharge summaries and death certificates, capturing both kidney-related events and mortality. *N*-glycomic analysis was performed using UHPLC using the same experimental conditions as described for the other studies. The gathered data was used to examine the association of IgG glycans with prevalent diabetes mellitus (either type 1 or type 2 diabetes mellitus; N=1868 / 35.8 %; DM1 study), cardiovascular disease (defined as prevalent coronary heart disease, history of stroke or prevalent peripheral arterial disease; N=1591 / 30.5%; CVD1 study) and all-cause mortality. The GCKD study is registered as a national clinical study (DRKS 00003971) and was approved by local ethics review boards of all participating institutions. The study adheres to the principles set out in the Declaration of Helsinki

#### PRECISESADS

The PRECISESADS cohort included a total of 854 serum samples provided by collaborators from the 3TR consortium. 542 originated from cross-sectional collections and 59 from an inception cohort, while 263 samples represented healthy control subjects. After quality control, 92 healthy control samples and 88 rheumatoid arthritis (RA3), 128 Sjögren’s disease (SjD1) and 87 systemic sclerosis (SSc) samples were included in the analysis. All data were fully anonymized prior to transfer, and available metadata included only age, sex, and diagnostic category. The samples were previously characterized according to clinical diagnosis and stratified into molecular clusters as described in the PRECISESADS study^50^. Plasma IgG was isolated and analyzed for *N*-glycan composition using UHPLC under the same experimental conditions as for the other studies. The study adhered to the standards set by International Conference on Harmonization and Good Clinical Practice (ICH-GCP), and to the ethical principles that have their origin in the Declaration of Helsinki (2013). Each patient signed an informed consent prior to study inclusion. The Ethical Review Boards of the participating institutions approved the protocol.

#### EZE-DZHK

The EZE-DZHK cohort represents a cross-sectional, multi-disease study encompassing a spectrum of chronic inflammatory conditions, established at the Christian-Albrechts-University of Kiel (CAU), Germany, within the framework of the broader SYSCID (Systems Medicine of Chronic Inflammatory Diseases) initiative. This cohort was designed to capture shared and disease-specific molecular features across immune-mediated disorders and includes patients sampled at a single clinical time point under standardized conditions. The study population comprised a total of 871 individuals, including patients diagnosed with five distinct chronic inflammatory diseases (EZE): systemic lupus erythematosus, ulcerative colitis, Crohn’s disease, rheumatoid arthritis, and psoriatic arthritis, as well as a control group consisting of individuals with cardiovascular, non-inflammatory conditions (DZHK). After quality control, a total of 609 samples were retained for downstream analyses. The final dataset included 197 control samples, 52 SLE samples (SLE4 study), 71 UC samples (UC4 study), 99 CD samples (CD4 study), 87 RA samples (RA2 study) and 103 PsA samples (PsA1 study). Total plasma IgG was isolated using standardized purification protocols to ensure comparability across disease groups. Subsequent characterization of IgG *N*-glycosylation was performed at the subclass level, employing subclass-specific Fc glycopeptide analysis following tryptic digestion. Glycopeptides were analyzed using LC-MS as described in the subsequent section. All study procedures were conducted in accordance with the ethical principles outlined by the SYSCID consortium and adhered to relevant institutional and regulatory guidelines governing human subject research. The study was conducted in accordance with the Declaration of Helsinki and approved by the ethics committee of the Christian-Albrechts-Universität zu Kiel (A 124/14).

#### Hormone replacement therapy

To evaluate the effect of HRT on IgG glycans, female patients from the Newson Clinic who initiated HRT at the time of recruitment were selected. Patients were started on oestrogen in combination with progesterone, testosterone or both (Supplementary Table 8). The analysis included 19 women, followed for up to 15 months (mean follow-up = 10.1 months). The mean age at baseline was 52.6 years (sd = 4.15). Participants underwent repeated measurements during the study period: five women had two measurements, eleven had three measurements, and three had four measurements. Detailed information about the study characteristics can be found in Supplementary Table 8 and Supplementary Table 9. At each time point the dried blood spots were collected for the IgG *N*-glycome measurements that were performed using UHPLC under the same experimental conditions as for the other studies. The study protocol was approved by the Genos ethic committee and all participants signed the written consent.

#### Therapeutic plasma exchange

To investigate the effects of TPE on IgG glycans, nine participants who underwent TPE were recruited. The intervention was administered over a period of up to six consecutive months. The participants consisted of four females and five males with the mean age of 60.8 years (sd = 9.19). The dried blood spots for IgG *N*-glycome measurements were collected three times: at baseline, at average of 2.99 months for second time point and at the end of the study with mean maximum duration of 5.05 months. Detailed information about the study characteristics can be found in Supplementary Table 9 and Supplementary Table 11. IgG glycome was profiled using CGE as described in the following paragraphs. The study protocol was approved by the Diagnostics Institutional Review Board (Cummaquid, MA). All participants provided written informed consent.

### IgG *N*-glycan profiling

Details of study-specific pre-analytical procedures are provided in the original studies referenced in Supplementary table 1 (Reference column). The analytical workflows applied across cohorts followed a general approach, which is summarized below to provide an overview of how IgG glycosylation was profiled, while exact methodological implementations can be found in the respective publications.

#### UHPLC-based IgG *N*-glycan profiling

For samples analyzed by UHPLC, IgG was isolated from plasma or serum using Protein G affinity chromatography, as previously described by Pučić et. al.^51^ and Trbojević-Akmačić *et al.*^52^. The isolated IgG underwent enzymatic release of *N*-glycans using PNGase F fluorescent labelling with 2-aminobenzamide (2-AB). Excess reagents were removed using solid-phase extraction, yielding labelled glycans suitable for chromatographic analysis. Separation of glycans was performed by hydrophilic interaction ultra-high-performance liquid chromatography (HILIC-UHPLC), allowing resolution of major IgG glycan species into discrete peaks. While column types and instrument parameters varied between studies, chromatograms were processed and curated to ensure consistent integration. The identities of chromatographic peaks had been previously confirmed by LC–MS analysis in earlier studies^51,53^. This approach enables robust and high-throughput quantitative profiling of IgG *N*-glycans and allows for cross-study comparisons after data harmonization. For PRECISESADS studies, an adjusted UHPLC protocol using RapiFluor-MS labeling was applied. IgG was isolated from plasma or serum using CIM^®^ r-Protein G LLD 0.2 mL Monolithic 96-well Plate (2 µm channels) (Sartorius BIA Separations, Slovenia) as previously described by Trbojević-Akmačić et. al.^52^. Approximately 15 μg of IgG was dried under vacuum in PCR plates. Subsequent deglycosylation, fluorescent labeling, and cleanup were performed using the GlycoWorks™ RapiFluor-MS *N*-Glycan Kit (Waters Corporation, USA) following the manufacturer’s protocol (Waters Corporation 2017) and Deriš et al.^54^. Briefly, dried IgG was resuspended in 5% (w/v) RapiGest SF, denatured at 99 °C for 3 min, and deglycosylated with PNGase F at 50 °C for 5 min. Released *N*-glycans were labeled with RapiFluor-MS reagent for 5 min at room temperature, followed by HILIC-SPE cleanup using GlycoWorks μElution plates. Labeled glycans were analyzed by HILIC-UHPLC with fluorescence detection, and peaks were integrated and quantified as relative (% area) values.

#### CGE-based IgG *N*-glycan profiling

For the TPE study, IgG glycome profiling was performed using capillary gel electrophoresis with laser induced fluorescence (CGE-LIF) analysis following the protocol described by Hanić et al.^55^, which builds upon previously published methods by Pučić et al.^51^ and Trbojević-Akmačić et al.^52^. Briefly, the procedure involved IgG isolation, followed by glycan release with PNGase F, fluorescent labelling with APTS, and subsequent glycan cleanup using wwPTFE filter plate with Bio-Gel P-10. Labelled glycans were analysed on an ABI 3500 Genetic Analyzer (Thermo Fisher Scientific, USA), and electropherograms were annotated and integrated as described by Hanić et al.^55^.

#### LC–MS–based IgG Fc glycopeptide profiling

For samples analyzed by LC–MS, IgG was purified from plasma samples by Protein G affinity chromatography, as described previously^51,52^. Plasma was clarified by centrifugation and filtration before dilution in phosphate-buffered saline (PBS) and application to protein G monolithic plates. After repeated washing with PBS, IgG was eluted with formic acid and neutralized with ammonium bicarbonate. Purified IgG was digested overnight with sequencing-grade trypsin to generate Fc glycopeptides, which were subsequently cleaned and enriched using reversed-phase solid-phase extraction. The resulting glycopeptide mixtures were dried, reconstituted, and analyzed by nanoLC separation on Waters systems coupled to high-resolution Bruker quadrupole time-of-flight mass spectrometers equipped with nanospray ionization. Spectra were acquired in positive ion mode, and glycopeptides were annotated on the basis of accurate mass, diagnostic oxonium ions, and reference data from prior studies.

### Data processing and statistical analysis

To remove experimental variation from measurements normalization and batch correction were performed, on each UHPLC/LC-MS dataset separately. For UHPLC datasets, normalization by the total area was performed where the peak area of each of 24 (or 22 in case of RapiFluor-MS labeling) glycan structures was divided by the total area of the corresponding chromatogram. For LC-MS datasets, signals of interest were normalized to the total area of each IgG subclass. For LC-MS datasets, in further analyses only IgG1 subclass glycopeptides were used. Prior to batch correction, normalized glycan and glycopeptide measurements were log-transformed due to the right-skewness of their distributions and the multiplicative nature of batch effects. For both UHPLC and LC-MS datasets, the batch correction was performed on Iog-transformed measurements using the “ComBat” method (R package sva) where the technical source of variation was modelled as a batch covariate. To get measurements corrected for experimental noise, estimated batch effects were subtracted from log-transformed measurements. Seven derived traits were calculated from glycan/glycopeptide measurements directly obtained by UHPLC/LC-MS analyses – total agalactosylated glycans (G0), total monogalactosylated glycans (G1), total digalactosylated glycans (G2), total galactosylated glycans (G), total sialylated glycans (S), total fucosylated glycans (F) and total glycans with bisecting GlcNAc (B), as described before^51^. These derived traits average glycosylation features across different individual glycan structures and are consequently more closely related to individual enzymatic activities and underlying genetic polymorphisms. Glycan age was calculated for each dataset separately by implementing general regression model where G0, G2 and S were used as predictors, chronological age was used as response variable, and model prediction was defined as glycan age. Differences in *N*-glycosylation of IgG between cases and their respective controls were analyzed using a general linear model. Age and sex variables were included in the model to control for their effects. Only in 4 aging cohorts, instead of case/control status, chronological age (expressed in decades) was included in regression model as main predictor. Regression analyses were performed for each cohort separately. Prior to analyses, glycan variables (except glycan age) were all transformed to standard Normal distribution (mean = 0, sd = 1) by inverse transformation of ranks to Normality (R package “GenABEL”, function rntransform). Using rank transformed variables in analyses makes estimated effects of different glycans in different studies comparable as transformed glycan variables have the same standardised variance. False discovery rate was controlled using the Benjamini–Hochberg procedure (function p.adjust (method = “BH”)). Data was analysed and visualised using R programming language (version 4.3.2). Associations between glycan age and mortality were assessed using Cox proportional hazards models, with survivors censored at the last follow-up. Analyses were restricted to a maximum follow-up of 8.5 years, with a median observed follow-up of the same duration. Model 1 was adjusted for age and sex. Model 2 was further adjusted for body mass index, smoking status, systolic blood pressure, LDL cholesterol, cardiovascular disease (peripheral arterial occlusive disease, coronary heart disease, or stroke), and diabetes (ATC A10 medication or HbA1c ≥6.5%). Model 3 was additionally adjusted for C-reactive protein, and Model 4 additionally included urinary albumin-to-creatinine ratio (UACR) and estimated glomerular filtration rate (eGFR), the latter defined as the mean of creatinine- and cystatin C–based equations from the European Kidney Function Consortium. eGFR, UACR, LDL cholesterol, and C-reactive protein were log-transformed. Of 5,217 participants, 5,086 had valid glycosylation measures; 4,827 had complete covariate data and were included in the analyses. The analysis was replicated in the Vis cohort^9,56^ with the same adjustments, excluding eGFR and UACR, with the 10-year follow up period. To investigate the relationship between menopausal status and glycan age linear mixed-effects model was fitted (lmerTest package)^57^. The model included menopausal status (MNPS) and age as fixed effects, with a random intercept for each cohort. To evaluate the influence of BMI on glycan age, a linear mixed-effects model was fitted using the lmerTest package in R^57^. The model included BMI, age and their interactions as fixed effects, with cohort ID modelled as a random intercept to account for cohort-level variability. Prior analysis BMI and age were centered by subtracting their means to avoid collinearity issues. The interaction term (age * BMI) allowed assessment of BMI effects across age strata. Estimated marginal trends of glycan age with respect to BMI were computed using the emmeans package^58^, stratified by age (in decades). Confidence intervals and p-values were reported for each trend estimate. To assess the effect of HRT, linear mixed-effects model was used where glycan age was modeled as dependent variable and time (number of months) was modeled as a fixed effect, participant ID was included as a random intercept, with chronological age included as additional covariate. To investigate the effect of TPE, changes in glycan age were assessed using linear mixed model where glycan age was modelled as dependent variable, time (in months) was modelled as fixed effect, ID of participant was included as random effect and age and sex were included as covariates. To assess the effect of caloric restriction, participant data from previously published dietary intervention study Diogenes^28^ were evaluated but this time focusing on the newly calculated glycan age. Analysis of longitudinal data for participants collected at baseline and after 8-week period was performed using linear mixed-effects model where glycan age was modeled as dependent variable and time was modeled as a fixed effect, participant ID was included as a random intercept, with age, sex, and BMI included as additional covariates. The analyses were first performed for each center separately and then combined using a random effects meta-analysis approach (R package meta, metagen (method.tau = “ML”; ML- maximum likelihood)).

### Data Availability

#### TwinsUK

The data used in this study are held by the Department of Twin Research at King’s College London. The data can be released to bona fide researchers using our normal procedures overseen by the Wellcome Trust and its guidelines as part of our core funding (https://twinsuk.ac.uk/resources-for-researchers/access-our-data/).

#### ORCADES

There is neither Research Ethics Committee approval, nor consent from individual participants, to permit open release of the individual level research data underlying this study. The datasets analysed during the current study are therefore available via managed access from, following approval by the Viking Genes Data Access Committee and in line with the consent given by participants. Each approved project is subject to a data or materials transfer agreement (D/MTA) or commercial contract.

#### HT study

The raw data are available on the PRIDE archive (http://www.ebi.ac.uk/pride) under the identifier: PXD015684.

Data from the other studies is available from the corresponding authors upon request.

## Acknowledgments

This work was supported by the Human Glycome Project. Equipment and products from GlycanAge, Waters and New England Biolabs®, Inc. were used for this research. IgG glycome analysis was supported through the following Horizon Europe grants: GlycanSwitch (contract no. 101071386), SYNHEALTH (contract no. 101159018) and AttractAdria (contract no. 101216942). TwinsUK is funded by the Wellcome Trust, Medical Research Council, Versus Arthritis, European Union Horizon 2020, Chronic Disease Research Foundation (CDRF), Wellcome Leap Dynamic Resilience Programme (co-funded by Temasek Trust), Zoe Ltd, the National Institute for Health and Care Research (NIHR) Clinical Research Network (CRN) and Biomedical Research Centre based at Guy’s and St Thomas’ NHS Foundation Trust in partnership with King’s College London. CM is funded by the Chronic Disease Research Foundation (CDRF) and by the Italian Ministry of Education and Research (MUR): Dipartimenti di Eccellenza Program 2023-2027. The Orkney Complex Disease Study (ORCADES) was supported by the Chief Scientist Office of the Scottish Government (CZB/4/276, CZB/4/710), a Royal Society URF to J.F.W., the MRC Human Genetics Unit quinquennial programme “QTL in Health and Disease”, Arthritis Research UK and the European Union framework program 6 EUROSPAN project (contract no. LSHG-CT-2006-018947). The GCKD study was and is supported by the BMBF (FKZ 01ER 0804, 01ER 0818, 01ER 0819, 01ER 0820 and 01ER 0821) and the KfH Foundation for Preventive Medicine. Unregistered grants to support the study were provided by corporate sponsors (listed at https://gckd.org). We are grateful for the willingness of the patients to participate in the GCKD study. The enormous effort of the study personnel of the various regional centers is highly appreciated. We thank the large number of nephrologists who provide routine care for the patients and collaborate with the GCKD study. The work of U.T.S. and A.K. was funded by German Research Foundation (DFG) project ID 431984000 (SFB 1453). The PRECISESADS studies received funding from the IMI 2 JU for the 3TR project (grant number 831434). The JU receives support from the EU Horizon 2020 research and innovation programme and EFPIA.

## Author contributions

These authors contributed equally: Anika Mijakovac, Elena Butz and Frano Vučković. G.L. and A.M. designed the study. Z.S.Y., J.Ć., F.Š., J.S.N., C.R.H.H., O.P., D.K., E.V., B.Y., L.N., C.M., C.J.S., T.D.S., J.H., E.P., M.S., M.T., J.F.W., M.E.A.R., K.A., P.R., A.F., N.F, S.S., D.M., A.K. and U.T.S. recruited the participants and provided plasma samples. B.R., H.D., B.R.T., F.L., A.C., I.G., N.Š.B., G.J., J.Š., I.B., J.K., T.P., M.H., M.P.B., I.T.A., T.Š., J.Š., M.M.K., M.F., A.C., M.P. and O.G. performed the glycan analyses. E.B., F.V., A.F.H. and D.K. performed the data analysis. A.M., E.B., F.V. and G.L. drafted the original manuscript. S.M.M., A.K., V.Z., D.P., U.T.S. and W.W. critically reviewed the study. All authors edited and approved the final version.

## ABBREVIATIONS

ADCC: Antibody-dependent cellular cytotoxicity
AF: Atrial fibrillation
AS: Allergic sensitization
BMI: Body mass index
CAD: Coronary artery disease
CD: Crohn’s disease
CGE: Capillary gel electrophoresis
CGE-LIF: Capillary gel electrophoresis with laser induced fluorescence
COPD: Chronic obstructive pulmonary disease
CRP: C reactive protein
CRC: Colorectal cancer
CVD: Cardiovascular disease
DM: Diabetes mellitus
eGFR: Estimated glomerular filtration rate
FNAIT: Fetal neonatal alloimmune thrombocytopenia
GlcNAc: *N*-acetylglucosamine
GVHD: Graft-versus-host disease
HF: Heart failure
HILIC-SPE: Hydrophilic interaction chromatography with solid phase extraction
HILIC-UHPLC: Hydrophilic interaction ultra-high-performance liquid chromatography
HRT: Hormone replacement therapy
HT: Hashimoto’s thyroiditis
IgG: Immunoglobulin G
LC-MS: Liquid chromatography coupled to mass spectrometry
LDL: Low-density lipoprotein
MS: Multiple sclerosis
PBS: phosphate buffer saline
PsA: Psoriatic arthritis
RA: Rheumatoid arthritis
SjD: Sjögren’s disease
SLE: Systemic lupus erythematosus
SSc: Systemic sclerosis
T2D: Type 2 diabetes
TPE: Therapeutic plasma exchange
UC: Ulcerative colitis
uACR: Urine albumin to creatinine ratio
UHPLC: Ultra-high-performance liquid chromatography

## Supplementary Figures

**Supplementary Figure 1.**
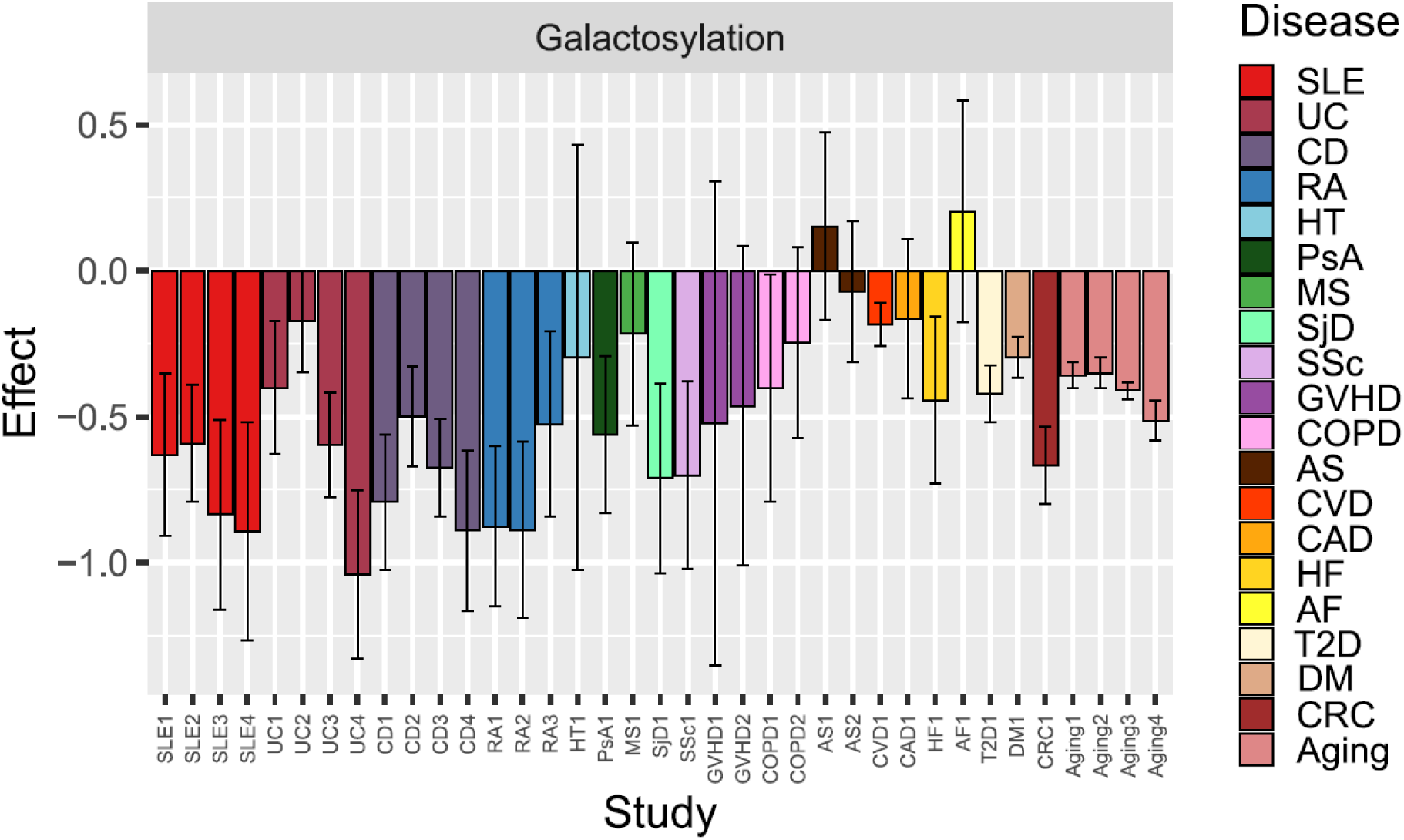
Total IgG galactosylation was calculated as a sum of total monogalactosylation (G1) and total digalactosylation (G2) obtained from glycan/glycopeptide measurements after UHPLC/LC-MS analyses. The X axis shows the individual studies representing the status of different diseases. The Y axis shows regression coefficients (β) obtained from linear models, representing the effect of disease status or chronological age on the total IgG galactosylation. For disease models, β represents the mean difference between cases and controls (in standard deviation units). For aging cohorts, β represents the change in total IgG galactosylation per 10-year increase in chronological age (SD units per decade). Models assessing disease status were adjusted for age and sex, whereas models assessing chronological age were adjusted for sex only. Total IgG galactosylation measurements were transformed to a standard normal distribution (mean = 0, SD = 1) using inverse rank-based normal transformation, enabling direct comparison of effect sizes across traits and studies.

**Supplementary Figure 2.**
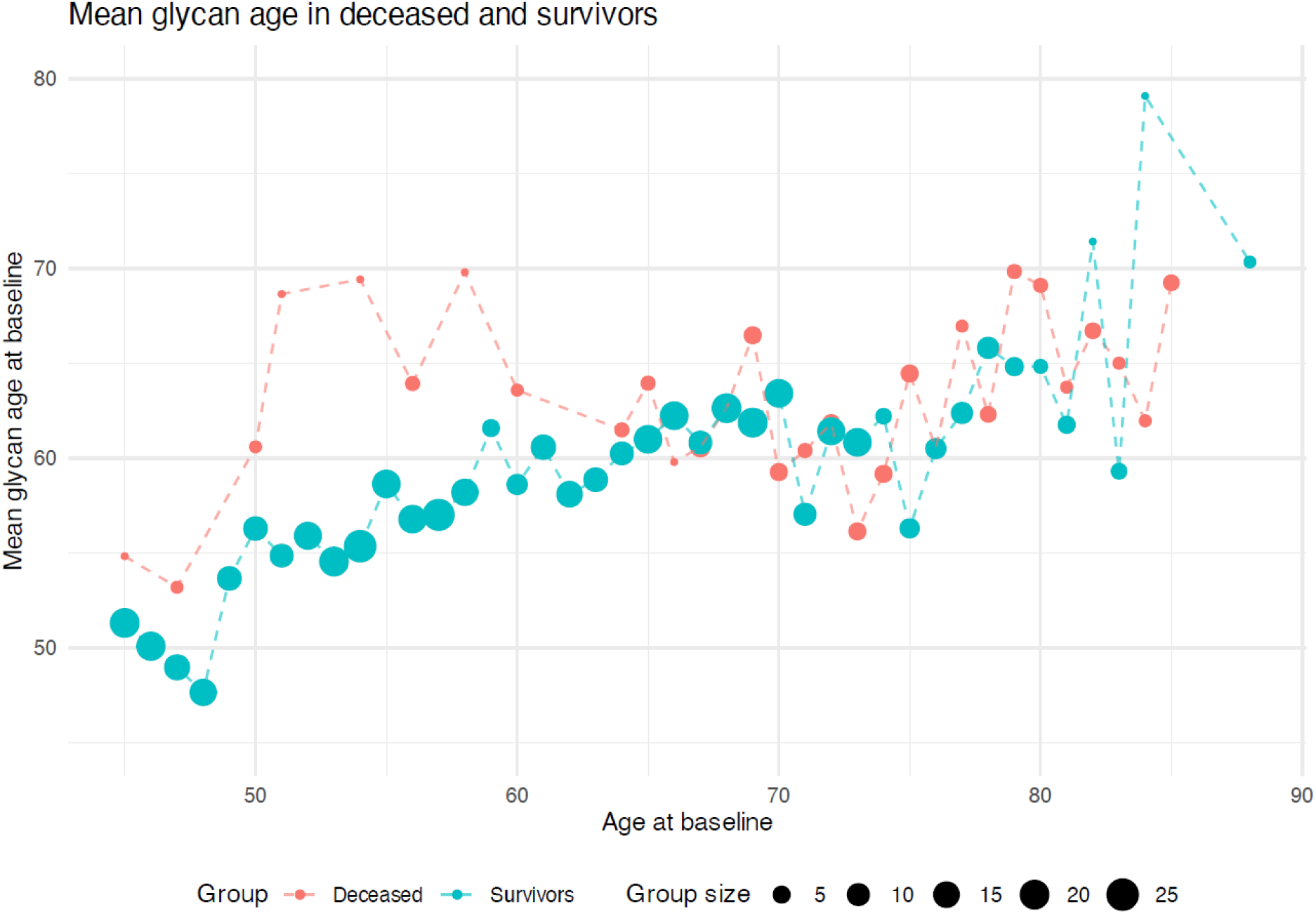
Mean glycan age in participants of the same chronological age who survived or died in the replication cohort (Vis). Y axis shows mean glycan age at the baseline visit for participants of the Vis study. X axis shows chronological age (in years) at the baseline visit of the Vis study for group sizes of 50 or more. Blue dots represent participants of the Vis study that did not die during the follow-up period of 10 years. Red dots represent the participants of the Vis study that died during the follow-up period due to any cause.

**Supplementary Figure 3.**
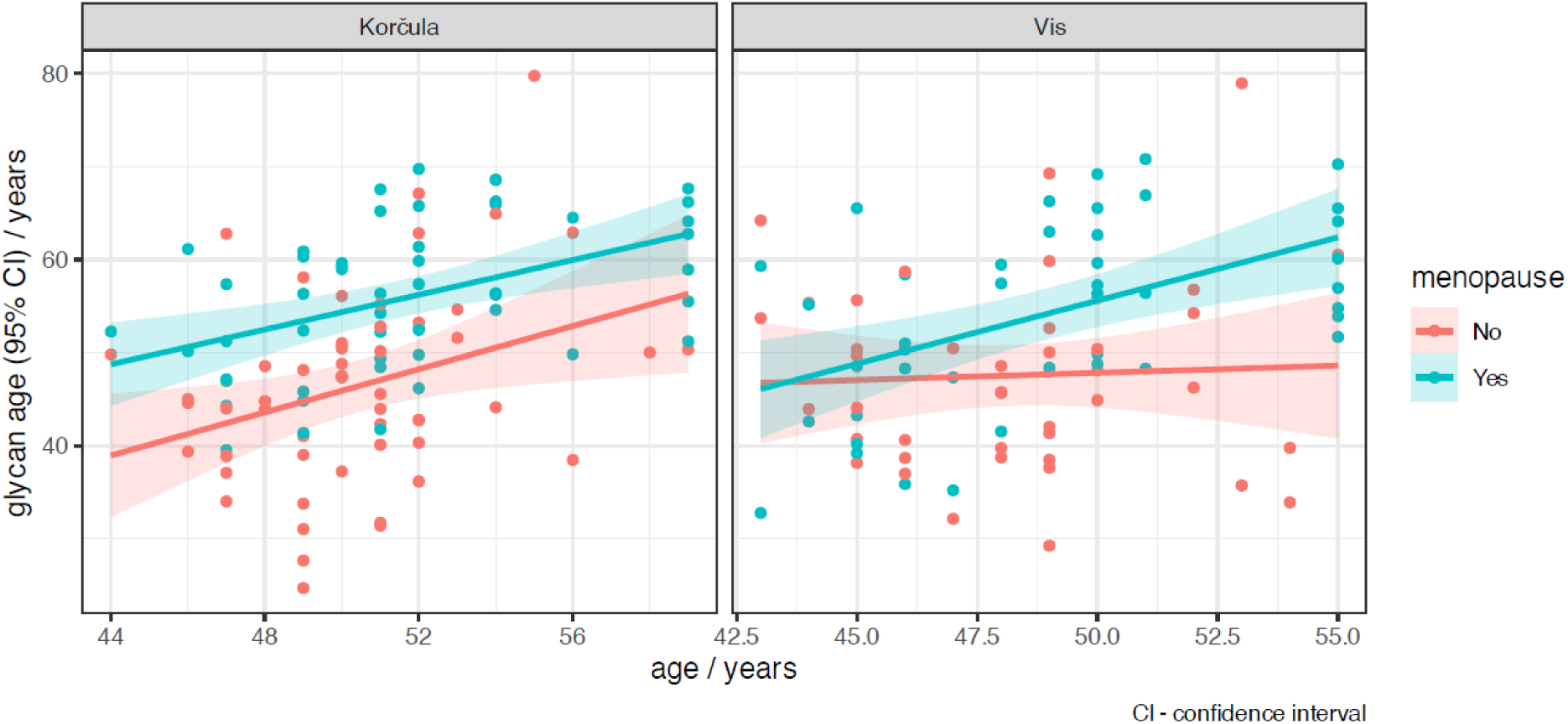
Mean glycan age of menopausal (blue) and premenopausal (red) women of the same chronological age in Korčula and Vis cohorts. Lines represent the mean levels of glycan age across chronological age. Shaded areas around the mean indicate 95% confidence interval (CI).

**Supplementary Figure 4.**
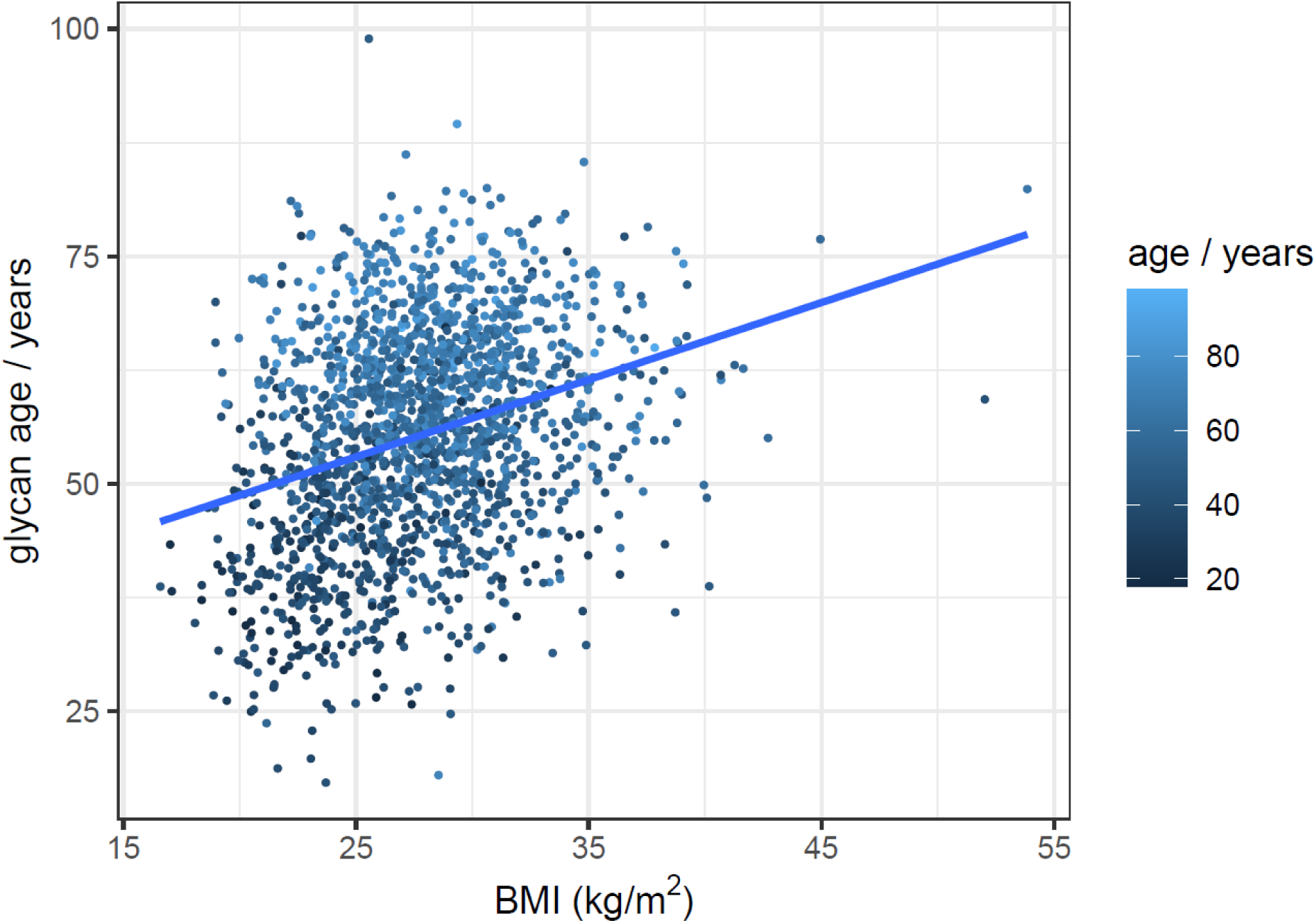
The association of glycan age and BMI in the Vis and Korčula cohorts. Points are colored by chronological age, with lower age depicted in dark blue and higher age depicted in light blue.

**Supplementary Figure 5.**
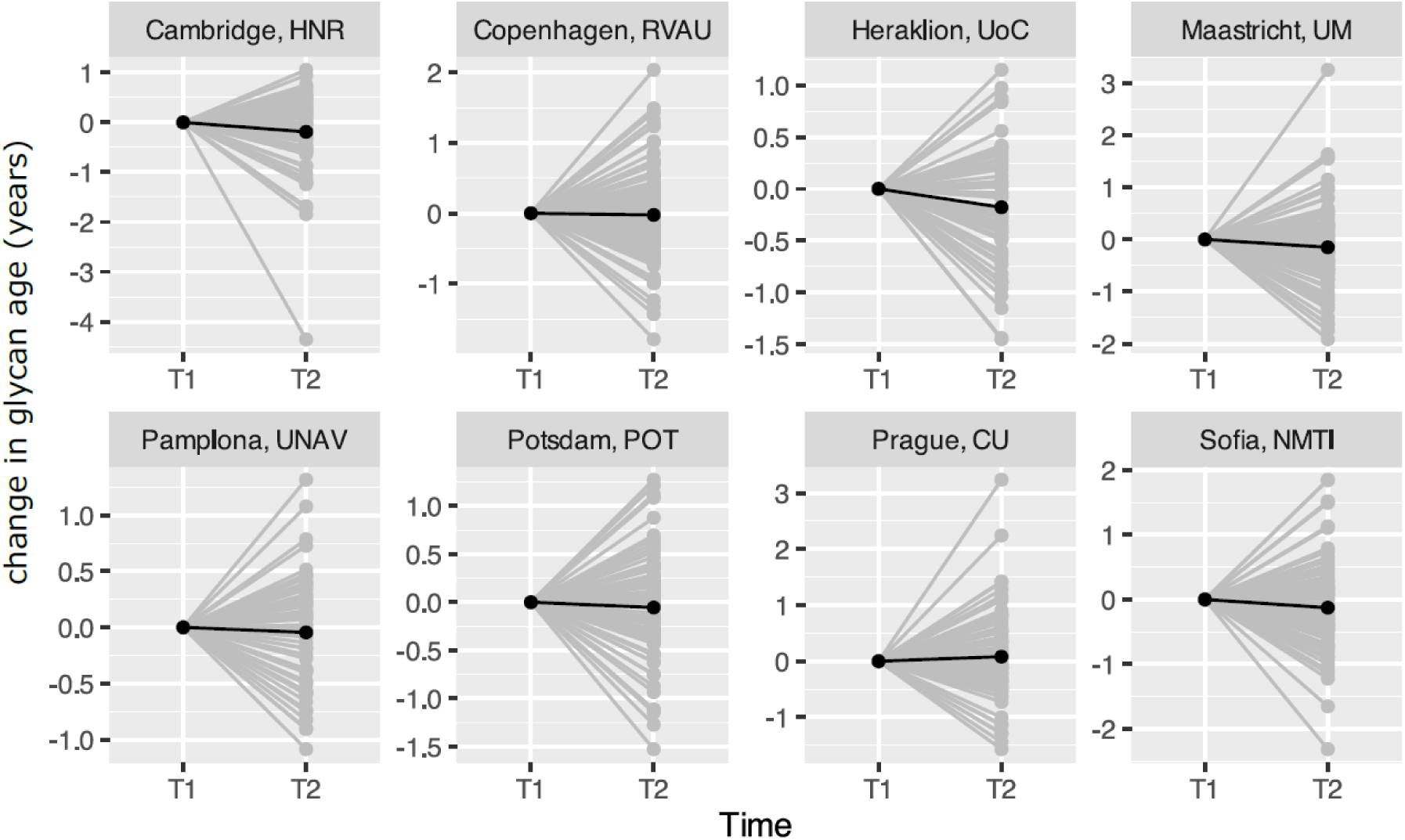
Change in glycan age from baseline across different centers after calorie restriction over 8 weeks period. T1- time point 1; T2- time point 2; Change in glycan agwas calculated as glycan age at T2 – glycan age at T1.

